# Non-genetic component of height as a surrogate marker for childhood socioeconomic position and its association with cardiovascular and brain health: results from HCHS/SOL

**DOI:** 10.64898/2026.04.08.26350438

**Authors:** Jee-Young Moon, Paola Filigrana, Linda C Gallo, Krista M Perreira, Jianwen Cai, Martha Daviglus, Lindsay E Fernández-Rhodes, Olga Garcia-Bedoya, Qibin Qi, Bharat Thyagarajan, Wassim Tarraf, Tao Wang, Robert Kaplan, Carmen R. Isasi

## Abstract

Childhood socioeconomic position (SEP) can have lifelong effects on health. Many studies have used adult height as a surrogate marker for early-life conditions. In this study, we derived the non-genetic component of height, calculated as the residual from sex-specific standardized height regressed on genetically predicted height, as a surrogate for childhood SEP, using data from the Hispanic Community Healthy Study/Study of Latinos (2008-2011). A positive residual would indicate favorable early-life conditions promoting growth, while a negative residual indicates early-life adversity that may stunt the development. The height residual was associated with early-life variables such as parental education, year of birth, US nativity and age at first migration to the US (50 states/DC), supporting the validity of height residual as a surrogate for early-life conditions. Furthermore, a height residual was positively associated with better cardiovascular health (CVH) and cognitive function among middle-aged and older adults. Interestingly, among <35 years old, the height residual was negatively associated with the “Life’s Essential 8” clinical CVH scores. These results suggest the non-genetic component of height as a surrogate for childhood environment, with predictive value for CVH and cognitive function.

## Introduction

Studies have consistently shown that lower childhood socio-economic position (SEP) is associated with adverse health outcomes later in life (1–5). Adult height, which becomes largely fixed after adolescence, reflects the cumulative effects of genetic and early life environment factors such as nutrition and disease (6). Thus, attained height has been widely used as an indicator of early-life conditions (7–14). Previously, using data from the Hispanic Community Health Study/Study of Latinos (HCHS/SOL), a longitudinal cohort of Hispanic/Latino adults from four U.S. field centers, we reported that economic hardship during both childhood and adolescence periods was associated with shorter height (10), possibly due to its links with poor nutrition and illnesses during infancy, childhood, and adolescence. In addition, other studies from low or middle-income countries have shown that childhood growth restriction is an important predictor of adult height (15). Similarly, in countries where malnutrition is common, stunting predicts adult short stature (16, 17). A previous study of Mexican immigrants and Mexican Americans born in the US, found shorter stature among those who were foreign-born, suggesting the possible impact of early life socio-economic and nutritional deprivation (18).

In addition to the effects of childhood environment, height has a strong genetic contribution (19), with estimated heritability ranging from 40% to 52% across ancestries (e.g., 52% for Europeans, 44% for Hispanics/Latinos). This, in turn, suggests that environmental factors are also important determinants of height. A recent adult height GWAS identified 12,111 independent genomic loci from a very large sample size of 5.4 million individuals from diverse ancestries (19). The heritability estimated from those regions explained more than 90% of the overall heritability in all ancestries. In the present study, we build from this literature to assess the non-genetic component of height as a proxy index of early childhood environment, providing a window into adverse environmental conditions decades in the past.

Adult height has been suggested as a predictor for various health outcomes in adulthood (20–25) such as better metabolic (26) and cardiovascular health (21), as well as better cognitive function, less cognitive impairment (20), and lower mortality (24). In HCHS/SOL, we also observed that taller height was associated with better cardiometabolic traits such as lower blood pressure and total cholesterol (22). Taken together, these findings suggest that the childhood environment may influence the development of chronic diseases later in life (1–5). In this paper, we expand this prior work to examine (1) whether the non-genetic component of height is a valid surrogate marker of early life socio-economic position and (2) it is positively related to adult cardiovascular health and cognitive function.

## Methods

### Study population and study flowchart

HCHS/SOL is an ongoing population-based cohort study of 16,415 Hispanic/Latino adults (aged 18-74 years) who were selected using a multi-stage probability sampling design from four US field centers located in metropolitan areas (Chicago, IL; Miami, FL; Bronx, NY; San Diego, CA) with representation of six major Hispanic/Latino heritage groups in the US (i.e., Mexican, Puerto Rican, Cuban, Dominican, Central or South American) (27, 28). In brief, at the baseline in-person examination (2008–2011), participants completed a comprehensive assessment of cardiometabolic risk or protective factors, questionnaires in their language of preference, and a blood draw for DNA extraction and other biomarkers. Participants aged 45 years or older completed assessments of cognitive function (n=9,690). Among 13,000 HCHS/SOL participants consented for genotyping, 346 participants were excluded from the current study because of missing height or genetic data, leaving an analytic sample of 12,654 participants. A subset of the sample, who participated in the HCHS/SOL Sociocultural Study (29), provided additional measures of childhood socio-economic environment (N = 4,146). The HCHS/SOL study and the Sociocultural Ancillary Study were approved by the institutional IRBs at each field center and the data coordinating center. All participants provided written informed consent. Study #07-1008 was approved by the University of North Carolina at Chapel Hill. **Figure S1** provides an overview of the study flowchart.

### Genotyping and height polygenic score (PGS)

Genotyping was done on the HCHS/SOL Custom 15041502 B3 SNP array (Illumina Omni 2.5M array plus ∼150,000 custom SNPs). Imputation was performed (30, 31) using SHAPEIT2 (version 2.r778) (32) for pre-phasing and IMPUTE2 (version 2.3.0) (33, 34) for genotype imputation, with reference to the worldwide 1000 Genome Project phase 3 panel. Principal components (PC) for population structure and kinship coefficients were estimated simultaneously (35, 36), and genetic-analysis groups (Cuban, Dominican, Puerto Rican, Mexican, Central American, and South American) were defined using the top five genetic PCs to ensure genetic homogeneity within self-identified Hispanic/Latino background (30, 31).

We calculated a height polygenic score (height_pgs_) based on the SBayesR-produced SNPs and weights for height PGS for Hispanics (455,180 participants from 11 studies) in the recent meta-analysis of height GWAS (19, 37, 38). In that study, Yengo et al. applied the SBayesR method to the Hispanic-specific GWAS summary statistics, which included the HCHS/SOL participants as part of the training data. The out-of-sample prediction accuracy of the Hispanic-specific height PGS was reported to be 15.4% (R^2^), adjusting for age, sex, and the top 10 PCs. In our study, 1,245,097 SNPs on chromosomes 1-22 were available for the height PGS calculation, after excluding 417 non-available or non-matching-allele SNPs.

### Non-genetic component of height (height_rez_) estimation

We aimed to derive a non-genetic component of height (height_rez_) as a surrogate marker of childhood environment conditions such as better nutrition, favorable socioeconomic status (SES), and/or social environment. First, as in Yengo et al. (19), a height in cm (measured with a wall stadiometer (SECA 222, Germany)) was standardized for each sex (mean (SD) 156.6 (6.5) cm for female; 169.7 (7.0) cm for male). Second, a prediction model for the standardized height was fitted using a linear mixed effects model with height_pgs_, sex and the top five PCs for population structure – to account for differences by ancestry proportions - (30) as fixed effects and genetic relatedness, household, and census block as random effects (model 1). To further adjust for generational difference in height (39) and aging-related height loss (40), we also fitted a prediction model including age and age^2^ additionally (model 2). Finally, we obtained two versions of height residuals with and without accounting for age (height_rez_ and height_rez-age_, respectively) by subtracting a fitted height estimate (only from fixed effects) from the standardized height: height_rez_ = height Z score – predicted height from model 1, and height_rez-age_ = height Z score – predicted height from model 2. A positive height residual indicates that a person grew taller than expected based on the prediction model while a negative residual indicates that a person grew shorter than expected. Note that the expected height represents the mean height predicted from the observed population-level associations with the included predictors (e.g., height_pgs_, PCs, sex, age), not the genetic height potential. The height residual accounting for age (height_rez-age_) was used in multivariable models with health outcomes. In addition, we estimated the narrow-sense heritability of height (quantifying additive genetic effects) based on the genetic relatedness (41), and the proportion of explained variance by height_pgs_.

### Childhood environmental conditions: parents’ education level, place of birth, age at migration to US, economic hardship, household utilities

Participants answered the highest grade/level of education of father and mother, separately. We categorized paternal and maternal education attainments into 4 groups – elementary or less, middle school, high school, and college or more. We also defined parental educational attainment as the highest level of paternal or maternal education. Participants self-reported whether they were born in the US (50 US states and DC) and if not, when they migrated first to the US mainland from a foreign country or US territory, which were categorized into four groups (US born, migrated in ages 0-12, 13-18, and 19+) (42). In addition, participants were asked for their self-identified Hispanic/Latino background. We also defined birth cohorts (1930-1949, 1950s, 1960s, 1970s, 1980-1999) based on the birth year.

In the Sociocultural Ancillary Study, participants were asked yes/no questions about experiencing material deprivation during childhood (did your family ever experience a period of time when they had trouble paying for their basic needs such as food, housing, medical care, and utilities?). They were also asked yes/no questions whether their homes had basic utilities (plumbing, electricity, phone, sewer/septic tank, respectively) when they were growing up.

### Health domain of Life’s Essential 8 CVH and cognitive function

As a summary measure of cardiometabolic risk factors, we created 8 cardiovascular health (CVH) metrics, largely grouped into two domains – health factors and health behaviors, according to the American Heart Association’s Life’s Essential 8, with separate scoring system for ≥20 years old and <20 years (43). In the health-factor domain of CVH, we scored each of 4 health factors (body mass index, blood lipids, blood glucose, blood pressure) by measurements at HCHS/SOL visit 1, where each component scores from 0 to 100 (better health), as detailed in Lloyd-Jones et al. (2019) (43). Briefly, body mass index CVH was quantified by BMI, blood lipids CVH by non-HDL cholesterol and lipid-lowering medication, blood glucose CVH by fasting blood glucose, HbA1c, or medication, and blood pressure CVH by systolic and diastolic blood pressure or medication. Then, we calculated an overall health-domain CVH score by averaging the four health factor scores, among those with complete information. Life’s Essential 8 CVH scores have been validated against all-cause and cardiovascular disease (CVD)-specific mortality, although BMI and blood lipid CVH scores demonstrated U-shaped associations (44, 45).

Participants ≥45 years at baseline (n=9,596) were administered a brief battery of neurocognitive tests in the participants’ preferred language during face-to-face interviews (46) – 1) Six-Item Screener (<u>SIS</u>) for mental status (47), 2) Brief Spanish-English Verbal Learning Test (B-SEVLT) for verbal learning (<u>B-SEVLT-sum</u>) and memory (<u>B-SEVLT-recall</u>) (46, 48), 3) abbreviated Word Fluency Test (<u>WF</u>) for verbal functioning(49), 4) Digit Symbol Substitution Test (<u>DSS</u>) of the Wechsler Adult Intelligence Scale–Revised for psychomotor speed and working memory (50). These five cognitive measures were standardized (z-score) by the mean and SD. A global cognition score was then calculated by averaging the standardized scores of four cognitive measures (excluding SIS), followed by standardization of this average.

### Covariates

Covariates including participants’ highest education, household income, health insurance, marital status, and language preference at examination (English, Spanish) were obtained from questionnaires. Health behavior variables were created according to AHA Life’s essential 8 metrics for the health behavior domain – diet, physical activity, nicotine exposure, sleep (43).

For behavior-domain CVH scores, diet CVH score was quantified using Dietary Approaches to Stop Hypertension (DASH) score (51), physical activity CVH by Global Physical Activity Questionnaire (52), nicotine exposure CVH by smoking and exposure, and sleep CVH by self-report of sleep duration, where each was scored from 0 to 100 (better behavior).

### Statistical analysis

Descriptive statistics of the study population were summarized by mean (standard error) for continuous variables and percentage (standard error) for categorical variables, accounting for the complex survey design (a multi-stage survey scheme and a sample weight calibrated to the 2010 US population census). The association between height_rez_ and childhood environmental conditions was examined by the weighted mean (SE) of height_rez_ according to childhood conditions, followed by a survey linear regression of height_rez_ as an outcome and childhood condition as an exposure. Age-stratified analyses were performed between height_rez_ and childhood environment. To examine the association between height_rez-age_ and adulthood health, a series of survey linear regression models were fitted on health-domain CVH score or cognitive function as an outcome and height_rez-age_ as a predictor, adjusting for a set of covariates. We considered three sets of covariates for the health-domain CVH and cognitive function models to add sequentially – (1) age, sex, field center, self-reported Hispanic/Latino background, US nativity and years living in the US, marital status, and language preference; (2) participants’ education, income, and health insurance; (3) health-domain CVH score (only for cognitive function) and four health behavior CVH scores (diet, physical activity, nicotine exposure, sleep). In addition, age-stratified and birth-cohort-stratified analyses were performed to examine associations of height_rez-age_ with health-domain CVH scores and cognitive function. An interaction by age group or birth-cohort was further assessed by including the corresponding interaction with height_rez-age_. As people tend to lose height during late-mid-life, a sensitivity analysis was performed after excluding people over 60 years old.

## Results

### Characteristics of the study population

The study population included 51.1% females with a mean age of 41.5 years (SE=0.3, range 18-74 years). Fourteen percent of the study population were born in 1930-1949, 16% in the 1950s, 22% in the 1960s, 21% in the 1970s, and 27% in 1980-1999 (**Table 1**). The majority (77%) were born outside the 50 US states and DC, 56% migrated first to the US states/DC when they were 19 years or more, 11% when aged 13-18 years, and 11% when aged 0-12 years. People reported that 34% of their parents completed elementary school education or less, 13% completed middle school education, 28% high school, and 26% of college or more. Among those with additional childhood socioeconomic data, 51% experienced economic hardship during childhood (0-18 years old), of whom 46% experienced hardship when they were 0-12 years old. Less than 20% of this study population did not have access to basic utilities (plumbing, sewer/septic tank, electricity, separately) at home during childhood (**Table 1**).

**Table 1.** Characteristics of study population, HCHS/SOL.

|  | Sample size | Weighted mean<br>[SE] or % (SE)* |
| --- | --- | --- |
| Age, years | 12654 | 41.5 [0.3] |
| Year of birth |  |  |
| 1930-1949 | 2041 | 13.7 (0.5) |
| 1950-59 | 3281 | 15.9 (0.4) |
| 1960-69 | 3409 | 22.2 (0.5) |
| 1970-79 | 1841 | 20.9 (0.7) |
| 1980-1999 | 2082 | 27.3 (0.8) |
| Age group, years |  |  |
| 18-24 | 1234 | 16.1 (0.6) |
| 25-34 | 1545 | 21.1 (0.7) |
| 35-44 | 2291 | 21.6 (0.6) |
| 45-54 | 3840 | 19.2 (0.5) |
| 55-64 | 2682 | 13 (0.4) |
| 65+ | 1062 | 9 (0.4) |
| Sex |  |  |
| Female | 7457 | 51.1 (0.7) |
| Male | 5197 | 48.9 (0.7) |
| Height (cm) - Male | 5197 | 170.7 [0.16] |
| Height (cm) - Female | 7457 | 157.3 [0.15] |
| Paternal education |  |  |
| Elementary | 4963 | 41.7 (0.9) |
| Middle | 1281 | 13.1 (0.5) |
| High | 2131 | 26 (0.7) |
| College | 1504 | 19.2 (0.8) |
| Maternal education |  |  |
| Elementary | 5622 | 41.5 (0.8) |
| Middle | 1507 | 13.8 (0.5) |
| High | 2430 | 27.9 (0.7) |
| College | 1406 | 16.9 (0.6) |
| Parental education |  |  |
| Elementary | 4836 | 34.1 (0.8) |
| Middle | 1501 | 12.6 (0.5) |
| High | 2619 | 27.6 (0.6) |
| College | 2285 | 25.8 (0.8) |
| US born (50 states/DC) and age at<br>migrated to the US |  |  |
| Migrated at age 19 yrs+ | 7920 | 55.7 (1) |
| Migrated at age 13-18 yrs | 1268 | 10.5 (0.4) |
| Migrated at age 12 yrs or less | 1151 | 10.6 (0.5) |
| US born (50 states/DC) | 2262 | 23.1 (0.9) |
| Self-reported Hispanic background |  |  |
| Dominican | 1194 | 10.1 (0.7) |
| Central American | 1345 | 7.3 (0.6) |
| Cuban | 2031 | 22.2 (1.8) |
| Mexican | 4650 | 35 (1.6) |
| Puerto Rican | 2176 | 16 (0.8) |
| South American | 836 | 5 (0.3) |
| More than one backgrounds | 396 | 4.3 (0.3) |
| Self-reported childhood economic hardship during childhood** |  |  |
| During 0-12 years old | 1987 | 46.3 (1.2) |
| During 13-18 years old | 1592 | 38.1 (1.2) |
| Both during 0-12 and 13-18 years old | 1444 | 33.8 (1.2) |
| Non-availability of basic utilities in the home during childhood** |  |  |
| No plumbing | 900 | 17.6 (0.9) |
| No sewer/septic tank | 846 | 17.4 (1.5) |
| No electricity | 523 | 9.6 (0.7) |
| No phone | 2112 | 42.8 (1.3) |

\*Values were estimated accounting for complex survey design of the study.
\*\*From the Sociocultural Ancillary Study (N=4,146), a subset of HCHS/SOL.

### Height heritability and height polygenic score (height_pgs_)

We estimated the narrow-sense heritability of height to be 53% (95% CI = 48.2-57.7%) in this Hispanic/Latino population, based on genetic relatedness and with adjustment for sex and first 5 PCs (41). Additionally adjusting for age and age^2^, the heritability estimate was 53.7% (95% CI=48.9-58.4%).

The height_pgs_ explained a substantial proportion of this heritability. Specifically, after adjusting for sex and 5 PCs, it explained 35.9% of the variance in height Z-scores, corresponding to 68% of the estimated narrow-sense heritability. With additional adjustment for age and age^2^, the variance explained by height_pgs_ increased to 39.2%, corresponding to 73% of the heritability. The full model, including height_pgs_, sex, and PCs, explained 41.5% of total variance in height, which increased to 48.6% with the inclusion of age, and age^2^.

### Association of the non-genetic component of height (height_rez_) with childhood socioeconomic conditions

Figure 1(A) **(Figure S2** for the mean height residual by categories) shows that younger generations had a greater height_rez_ compared with those born in 1930-1949. These findings may indicate an overall improvement in the overall childhood socioeconomic and nutritional environment for younger generations. US born individuals and those who relocated to the US at a younger age (0-12 years) had a higher height_rez_ than those who relocated to the US at an older age, which suggests exposures to better childhood socioeconomic conditions. A higher parental education was associated with a higher height_rez_. These associations were substantially attenuated when using height_rez-age_, another measure of the non-genetic component of height that accounts for age, suggesting birth cohort effects (Figure 1 **(B)).** When examined by age group, the trends between height_rez-age_, and both parental education and US born/migration age period remained generally consistent (**Figure S3**).

**Figure 1.**
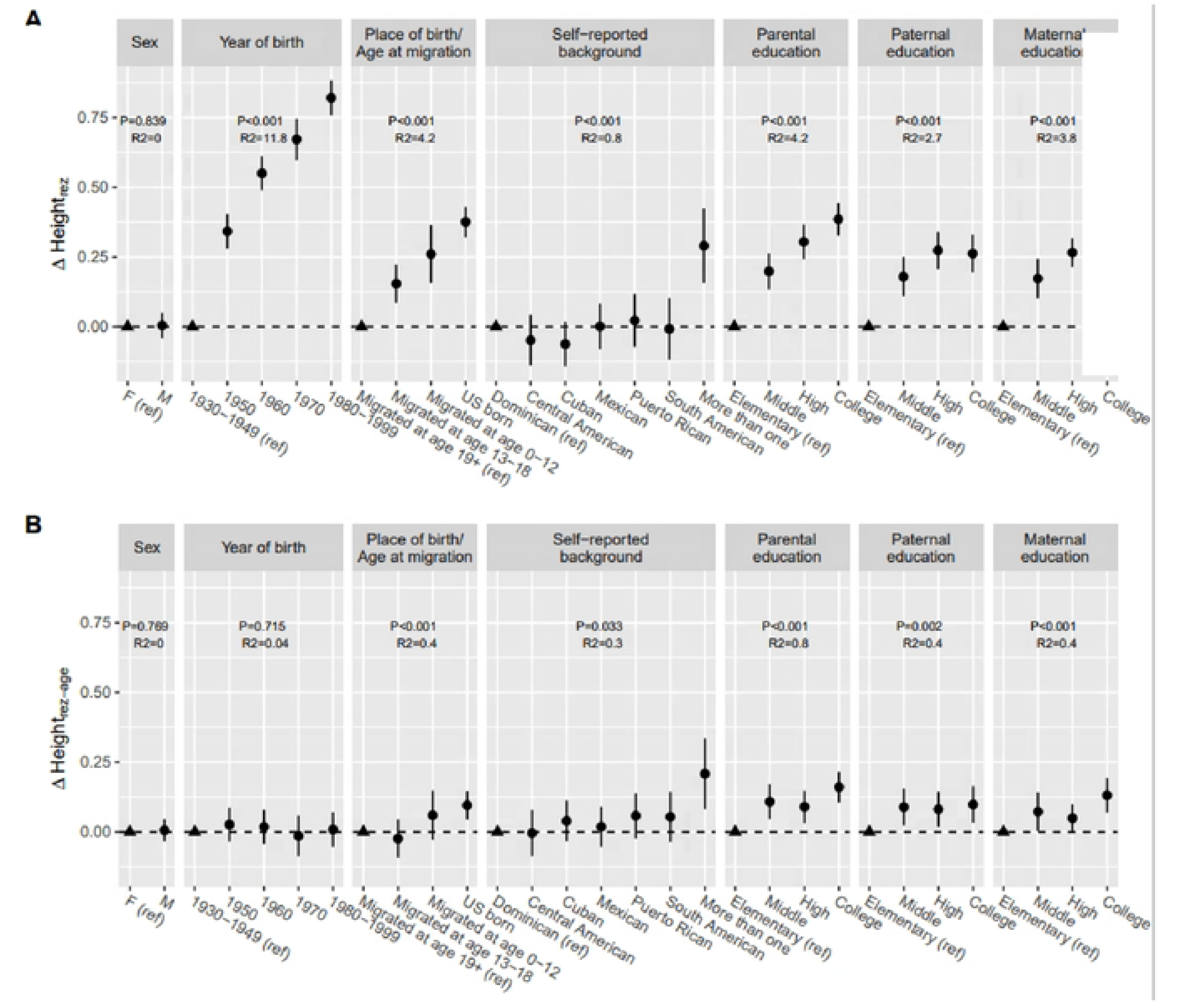
Difference in non-genetic component of height (Height_rez_, Height_rez-age_,) by socio-demographic factors. (A) Height_rez_: height residual by subtracting the predicted height by height PGS, sex, and 5 PCs from the sex-standardized height. (B) Height_rez-age_: height residual by subtracting the predicted height by height PGS, sex, 5 PCs, age, age^2^, from the sex-standardized height. A circle point represents the mean difference of height residual of a group compared to a reference group (ref: triangle point) and the error bar represents 95% CI of the mean difference, incorporating the complex survey. R^2^ represents an explained variance(%) of height_rez_ or height_rez-age_ by each variable.

Not having access to plumbing and electricity at home during childhood was negatively associated with both versions of height residuals (Figure 2), which indicates that lacking basic utilities during childhood is associated with an environment less supportive of growth and development. Economic hardship experienced during adolescence or during both adolescence and early/middle childhood was associated with shorter height residuals, but not statistically significant (Figure 2). No difference in height residuals was observed for hardship in early/middle childhood.

**Figure 2.**
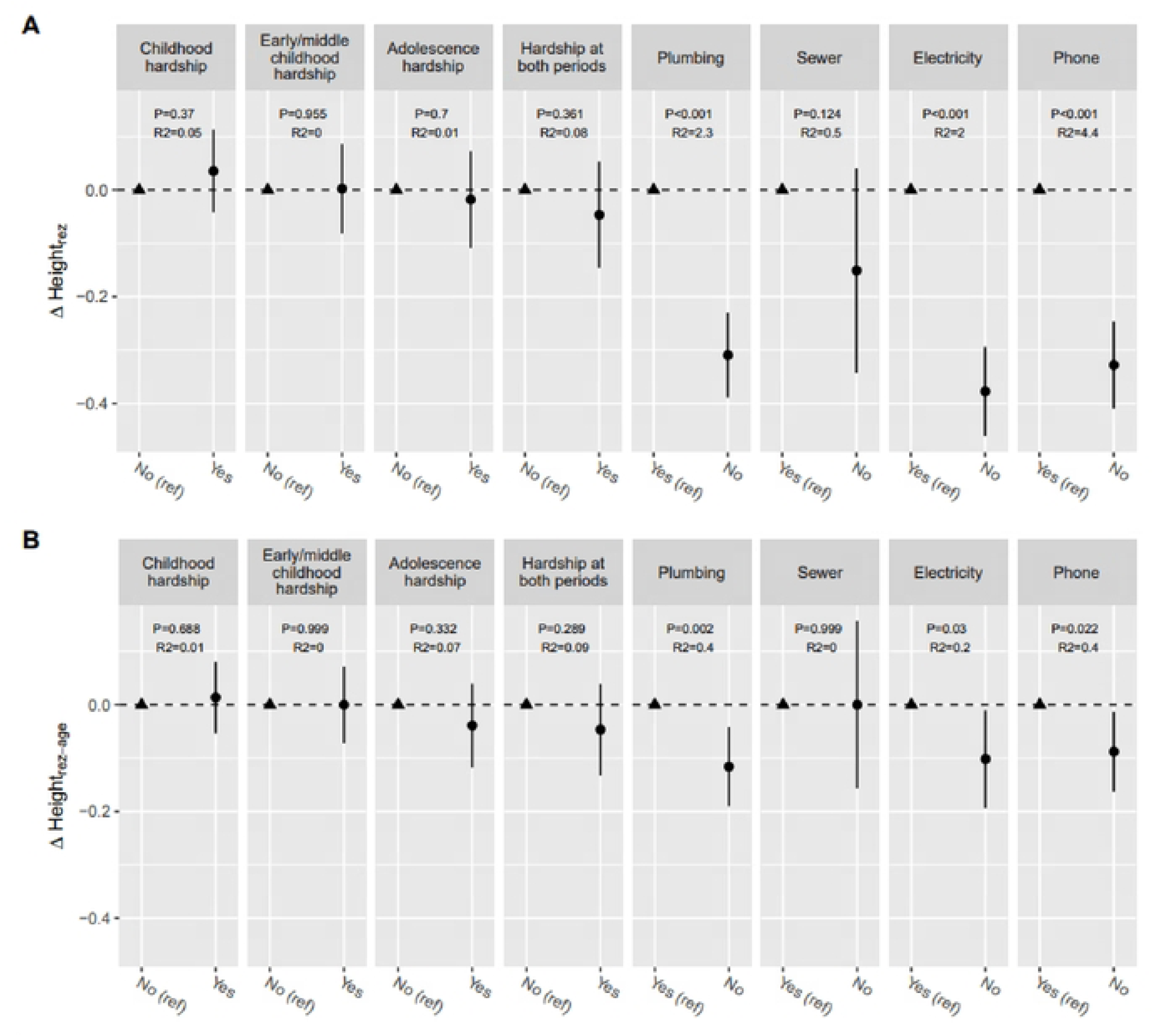
Difference in non-genetic component of height (Height_rez_, Height_rez-age_) by childhood economic hardship and basic utility availability from Sociocultural ancillary study (SCAS). (A) Height_rez_: height residual by subtracting the predicted height by height PGS, sex, and 5 PCs from the sex-standardized height. **(B)** Height_rez-age_: height residual by subtracting the predicted height by height PGS, sex, 5 PCs, age, age^2^, from the sex-standardized height. A circle point represents the mean difference of height residual of a group compared to a reference group (ref: triangle point), and the error bar represents 95% CI of the mean difference, incorporating the complex sutvey. R^2^ represents an explained variance(%) of height_rez_ or height_rez-age_ by each variable.

### Association of the non-genetic component of height (height_rez-age_) with CVH in adulthood

The clinical CVH score in our study population had a mean of 69.7 (SD=20.1). Mean (SD) values of each health factor CVH score were 85.7 (18.4) for glucose, 58.0 (20.0) for BMI, 72.7 (21.2) for blood pressure, and 62.3 (20.6) for non-HDL cholesterol. All component scores ranged from 0 to 100. Overall, height_rez-age_ was not associated with clinical CVH score. However, age-stratified analyses revealed heterogeneous associations across age groups (Figure 3). Among older groups (55-64 and 65+) there was a positive association between height_rez-age_ and clinical CVH score, suggesting that childhood socioeconomic environments more conducive to growth and development were associated with better adult cardiovascular health. In contrast, among younger individuals (18-24 and 25 -34 years old), strong inverse associations were observed with clinical CVH, BMI, blood pressure, and non-HDL cholesterol component scores, suggesting worse cardiometabolic profiles of young adults exposed to more advantaged early-life socioeconomic environments. Repeating the analyses stratified by birth cohort period identified similar patterns (Figure 4). Individuals born in more recent generations had worse clinical CVH, BMI and blood pressure scores when exposed to environments more conducive to growth and development, compared to the older generations. In addition, the use of the non-genetic component of height helped to separate out the effect of genetic component of height, as demonstrated in the association with blood pressure CVH score among individuals 45+ years (**Table S1**).

**Figure 3.**
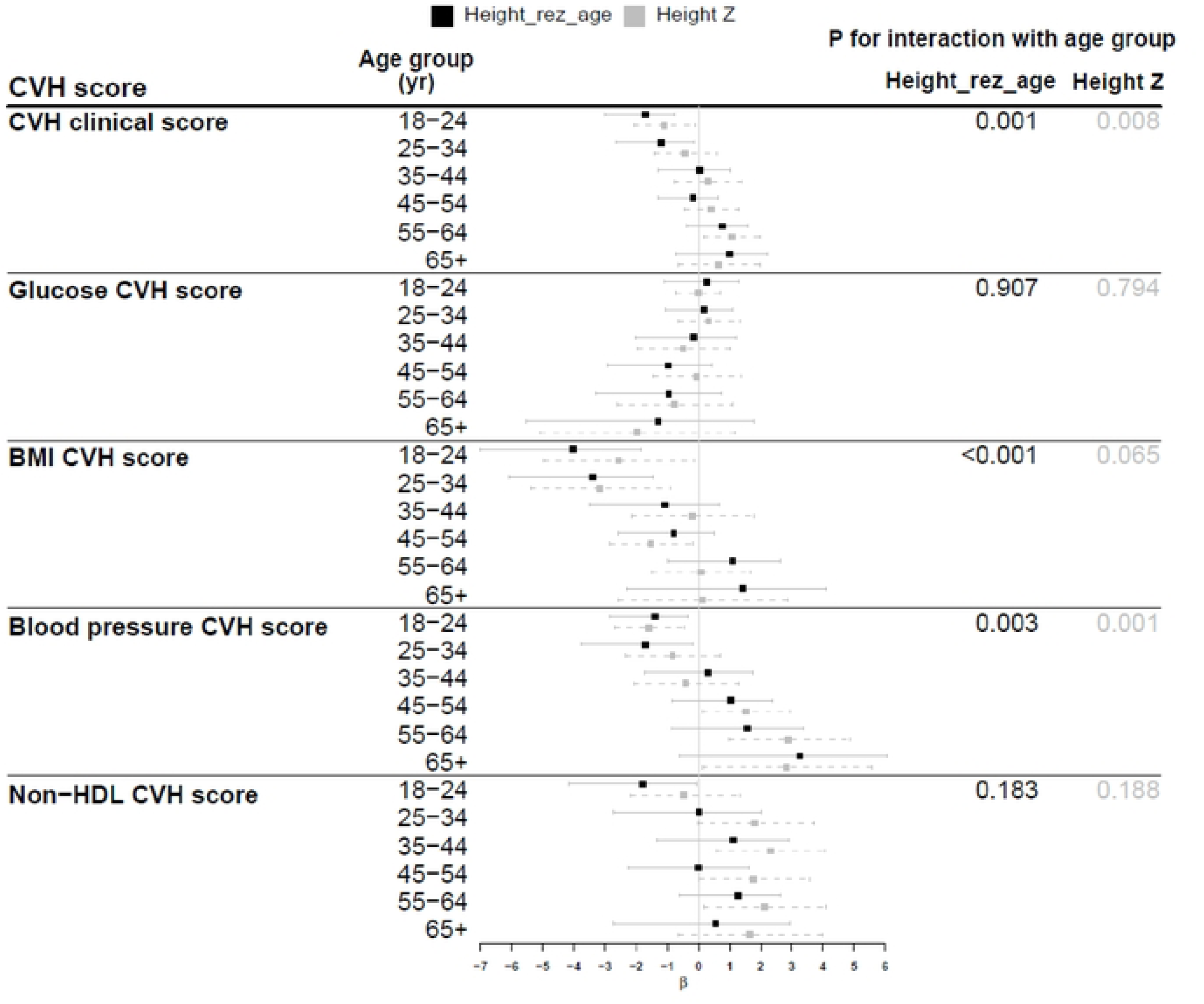
Association between the non-genetic component of height and health-domain CVB score by age groups. Height_rez-age_, is a height residual by subtracting predicted height by height PGS, sex, 5 PCs, age, and age^2^ from sex-stratified standardized height Z-score. Analysis is adjusted for age, sex, center, self-reported Hispanic background, US born, years in living in US, marital status, language preference, participant’s education, income, and health insurance, and AHA’s behavior CVH scores (diet, nicotine exposure, physical activity, sleep), accounting for complex survey design.pis per I SD of height_rez-age_ or height Z score.

**Figure 4.**
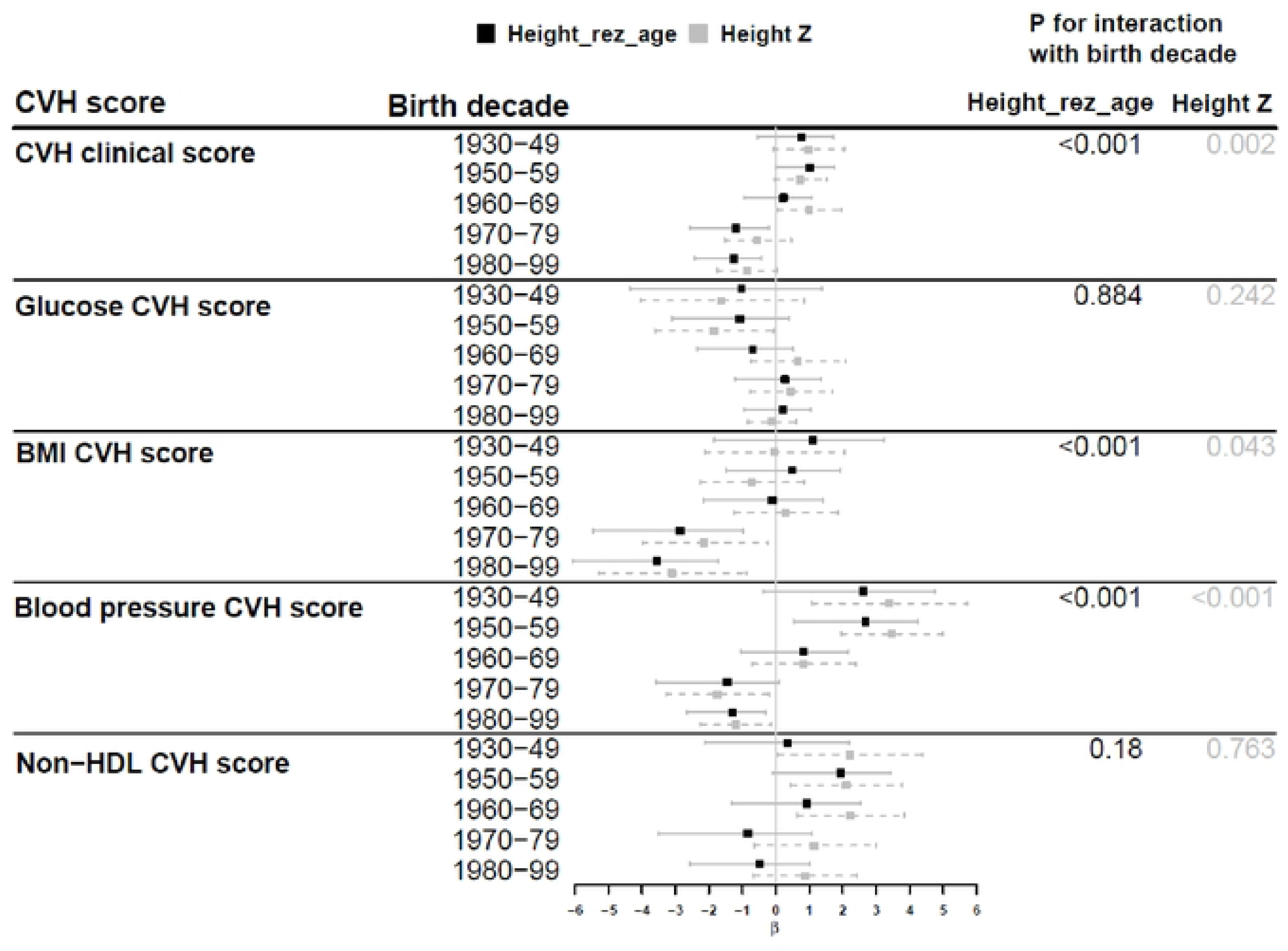
**Association between the non-genetic component of height and clinical-domain CVB score by birth year. Height_rez-age_** is a height residual by subtracting predicted height by height PGS, sex, and S PCs from sex-stratified standardized height Z-score. Analysis is adjusted for age, sex, center, self-reported Hispanic background, US born, years in living in US, marital status, language preference, participant’s education, income, and health insurance, and AHA’s behavior CVH scores (diet, nicotine exposure, physical activity, sleep), accounting for complex survey design. *β* is per I SD of height_rez_age_ or height Z score.

### Association of the non-genetic component of height (height_rez-age_) with cognitive function

Among persons aged 45+ years who completed assessments of cognitive function, we observed that a childhood environment more conducive to growth and development as indicated by a higher height_rez-age_ was associated with higher levels of cognitive function (**Table 2**). This was observed with measures of overall cognitive function (global cognition) and with each of the individual domains tested, except for the 6-item screener. In the stratified analyses, results were not materially different by age group or birth cohort (**Tables S2 and S3**). In addition, using height Z-score, as an alternative for childhood environmental conditions, resulted in similar positive associations with global cognition and each individual cognitive domain, albeit associations were slightly weaker (effect size) with wider confidence intervals than height_rez-age._

**Table 2.** Associations of the non-genetic component of height, measured height, and height PGS with cognitive function, among individuals aged 45+ years from HCHS/SOL.

| Outcome | Height<sub>rez-age</sub> |  |  | Height Z |  |  | Height PGS |  |  |
| --- | --- | --- | --- | --- | --- | --- | --- | --- | --- |
|  | Beta | 95% CI | R<sup>2</sup> (%) | Beta | 95% CI | R<sup>2</sup> (%) | Beta | 95% CI | R<sup>2</sup> (%) |
| Global cognitive score | 0.055 | 0.027, 0.082 | 0.4% | 0.046 | 0.013, 0.079 | 0.3% | 0.006 | -0.034, 0.047 | 0.0% |
| 6 item screener | 0.021 | -0.013, 0.055 | 0.0% | 0.034 | -0.004, 0.072 | 0.1% | 0.044 | 0.001, 0.087 | 0.1% |
| Spanish English verbal learning test - sum | 0.038 | 0.009, 0.067 | 0.2% | 0.040 | 0.006, 0.075 | 0.2% | 0.023 | -0.021, 0.066 | 0.0% |
| Spanish English verbal learning test - recall | 0.028 | 0.000, 0.055 | 0.1% | 0.009 | -0.023, 0.041 | 0.0% | -0.010 | -0.048, 0.028 | 0.0% |
| Word frequency | 0.046 | 0.009, 0.083 | 0.2% | 0.036 | -0.001, 0.073 | 0.1% | 0.000 | -0.044, 0.043 | 0.0% |
| Digit symbol substitution | 0.055 | 0.031, 0.080 | 0.5% | 0.056 | 0.028, 0.084 | 0.5% | 0.016 | -0.016, 0.048 | 0.0% |

Beta is cognitive function Z-score per 1 SD of height<sub>rez-age</sub>, height Z score, or height PGS.
R<sup>2</sup> (%) is an explained variance of outcome by height<sub>rez-age</sub>, height Z score, or height PGS.
A survey linear regression model was fitted for each cognitive function with height<sub>rez-age</sub> or height, accounting for complex survey design.
The model was adjusted for age, age<sup>2</sup>, sex, center, self-reported Hispanic background, US born, years in living in US, marital status, language preference, participant's education, income, health insurance, AHA's CVH clinical-score and four behavior CVH scores (diet, nicotine exposure, physical activity, sleep).

### Sensitivity analysis

To minimize a potential bias due to height loss in older individuals in deriving the height residuals, we conducted a sensitivity analysis restricted to individuals younger than 60 years. The non-genetic component of height (height_rez-age_) estimated in this group showed similar associations with childhood environmental variables (Figure S4), cardiovascular health indicators (Figure S5), and cognitive function (Table S4) as observed in the main analyses.

## Discussion

This study aimed to evaluate the discrepancy between genetically determined height and actual height, the non-genetic component of height, as a surrogate marker for early childhood socioeconomic environment. To our knowledge, there is very limited data on using the height residual to infer early life socioeconomic conditions (53). Consistent with our hypotheses we found that the non-genetic component of height was associated with indicators of early childhood socioeconomic environments, including parental education, US nativity, age at migration to the US 50 states/DC, and access to sanitation during childhood. The study findings suggest that individuals who were born in the US or migrated to the US at an early age (0-12 years old) were exposed to more favorable socioeconomic conditions that promoted optimal growth and development, resulting in taller non-genetic statures than the others. Similarly, our results showed that higher parental education (a common proxy for childhood socioeconomic status) was also associated with attaining taller stature relative to the genetically predicted height, providing additional evidence that height_rez_ could be used to infer about childhood nutritional environments. The observed association between height_rez_ and sanitation conditions (access to plumbing) at home during childhood also suggests that this biomarker is a useful indicator for environmental conditions related to socioeconomic status during childhood.

The study findings are consistent with prior literature that relates childhood socioeconomic status to child growth patterns. Data from the French EDEN mother-child cohort showed that low maternal education was associated with lower birth length and shorter stature from birth to 5 years (54), indicating that lower SEP influences growth patterns in children even in high-income countries, possibly due to suboptimal nutrition. Our study also shows that individuals who were U.S.-born or moved to the US at an early age (0-12 years old) were taller than individuals born abroad or relocated to the US at an older age. These findings are consistent with a previous study, showing shorter statures for Mexican non-migrants and Mexican immigrants to the US than US-born Mexican Americans (18). These findings further suggest that the non-genetic component of height reflects the potential influence of nutritional deprivation in a society.

Interestingly, the study showed birth cohort effects. Younger birth cohorts exhibited a higher non-genetic component of height than older birth cohorts, indicating more favorable growth conditions. This pattern could also be interpreted as younger birth cohorts attained stature closer to their genetic height potential – the genetically determined maximum height an individual can achieve - than older cohorts, with the latter experiencing greater stunting due to childhood malnutrition. However, it is important to note that our study used population-level height predictions based on genetic factors across the age spectrum, rather than estimating individual genetic height potential. In addition, the graded association of parental education with the non-genetic component of height was substantially attenuated when accounting for age. These secular trends in height have been widely reported worldwide as a reflection of changes in living conditions (55, 56) and nutritional environment (57). These trends of increasing statures have also been reported for Latin American countries. In Chile, for example, increasing height over 30 years was reported after the introduction of public health programs targeting malnutrition (58).

The birth cohort period effects were also observed when we examined the association of the non-genetic component of height with cardiovascular health indicators. A higher height residual, indicating an early-life environment promoting growth, predicted better cardiovascular health among middle-to-older aged individuals. However, the associations with CVH were reversed among those aged 18-35 years (born in 1980-2000). In the younger cohort, an early-life environment more conducive to growth (greater non-genetic component of height) was associated with poorer overall CVH, mostly driven by blood pressure and BMI. These counterintuitive findings may be explained by the obesogenic environment in which the younger generations grew up. The Framingham Heart Study reported a higher prevalence of obesity in offsprings compared to their parents at similar ages – 14.5% (BMI≥30kg/m^2^) in 1979-1983 for parents vs 30.4% in 1998-2001 for their offsprings (59). Our results are consistent to findings from Finnish cohorts evaluating the influence of height residual on coronary heart disease (CHD) risk (53). In these Finnish cohorts, the height residual was associated with SEP in the expected direction, and in turn, predicted CHD incidence. The authors also reported that birth year explained the variability of height, but they did not evaluate whether the associations of the height residual with CHD differed by birth cohorts.

Our study found that the non-genetic component of height was associated with better cognitive performance in midlife, suggesting enduring effects of early-life environments. Because cognitive function was assessed only among participants aged 45 years or older, we were unable to examine whether these associations extend to younger generations. Consistent with our results, other studies support the long-term influence of early growth on cognitive performance decades later. The study by Cohen-Manheim et al (60) in an Israel cohort demonstrated that higher stature in late adolescence and greater height gain from adolescence to young adulthood were both associated with better cognitive performance in midlife. In addition, a study in the UK biobank reported that genetic height was associated with better cognitive performance and reduced AD risk (61) – but this study did not assess the non-genetic contribution of height.

Although our findings support the use of the height residual as a surrogate marker of early life conditions - demonstrating its associations with health outcomes- we acknowledge limitations to these claims. Our study found small effect sizes and low R^2^ that are consistent with other studies. The Avon Longitudinal Study of Parents and Children (ALSPAC) reported small effects, with only a 0.8 cm difference in height residual between the most and least deprived neighborhood quintiles (62), compared with a 0.6 cm difference (corresponding to 0.12 SD) between the highest and lowest parental education levels in our study. In addition, economic hardship during adolescence or during both adolescence and early/middle childhood showed only a non-significant trend towards shorter height residuals in our study, and no difference was observed for hardship experienced during early/middle childhood. This pattern may be explained by delayed puberty among those exposed to childhood economic hardship, allowing a longer window for catch-up growth during adolescence (63). Alternatively, these null or weak associations may reflect lower accuracy of recalled family financial status (64), heterogeneity in economic conditions across Latin American countries, and limited sample size for these measures. The similar patterns observed between height Z-score and height residual in relation to health outcomes suggest that concerns of using height Z-score may be minimal, as environmental exposures are plausibly independent of genetic factors. Yet, in situations where the underlying genetic factors for height may confound the associations, the height residual offers a more valid approach.

Our study has several methodological limitations that need to be considered when interpreting results. While our study benefits from the increased power in detecting associated SNPs due to recent advances in the genetics of height, we acknowledge that there may still be unaccounted genetic factors influencing height in the Hispanic/Latino population (19). Although we attempted to consider intergenerational differences and height loss associated with aging (40) by incorporating age and age^2^ in the estimation of the height residual, it should be noted that individual variations in height loss may not be fully accounted for. Nonetheless, the sensitivity analysis supports the validity of our findings. Other limitations include the potential for residual confounding and the correlational – rather than causal – associations owing to the observational nature of the study. In addition, the study lacked prospectively collected measures of comprehensive early-life conditions such as nutrition and socioeconomic status. Further prospective studies following individuals from childhood to adulthood are needed to objectively validate the height residual against gold standard measures. Despite these limitations, the study offers an important glimpse of the long-lasting health effects of early life conditions. The negative association between height residual and CVH in young people suggests that the meaning of height as a biomarker for childhood environment and a predictor for health may change across time. Taken together, the study findings indicate that for adult cohorts that do not have direct measures of childhood exposures (e.g., EHR records with genetic data, All of US data), estimating the difference of attained vs. genetically predicted height is a good surrogate marker of early life environments.

## Data Availability

Data supporting results of this study from the HCHS/SOL are available in BIOLINCC at biolincc.nhlbi.nih.gov/requests/type/hchssol/. Researchers must be registered on the BIOLINCC website to access this form. Genetic data can be requested to HCHS/SOL at sites.cscc.unc.edu/hchs/

## Acknowledgements

This project was supported by the National Institute of Aging (RF1AG07761) and the Life Course Methodology Core (LCMC) at the New York Regional Center for Diabetes Translation Research (P30DK111022). The Hispanic Community Health Study/Study of Latinos was supported by contracts from the National Heart Lung and Blood Institute to the University of North Carolina (N01-HC65233), University of Miami (N01-HC65234), Albert Einstein College of Medicine (N01-HC65235), the University of Illinois at Chicago (HHSN268201300003I), Northwestern University (N01-HC65236), and San Diego State University (N01-HC65237). The following Institutes/Centers/Offices contributed to the HCHS/SOL through a transfer of funds to the National Heart Lung and Blood Institute: National Center on Minority Health and Health Disparities, the National Institute of Deafness and Other Communications Disorders, the National Institute of Dental and Craniofacial Research, the National Institute of Diabetes and Digestive and Kidney Diseases, the National Institute of Neurologic Disorders and Stroke, and the Office of Dietary Supplements.

## Conflict of interest

none declared.

### The contribution of authors to this study is summarized below

**Jee-Young Moon**: conceptualization, data curation, formal analysis, visualization, investigation, data interpretation, writing – original draft, writing – review & editing

**Paola Filigrana**: conceptualization, data curation, investigation, data interpretation, writing – review & editing

**Linda C Gallo**: investigation, data acquisition, funding acquisition, data interpretation, writing – review & editing

**Krista M Perreira**: data interpretation, writing – review & editing

**Jianwen Cai**: investigation, data acquisition, funding acquisition, data interpretation, writing – review & editing

**Martha Daviglus**: investigation, data acquisition, funding acquisition, data interpretation, writing – review & editing

**Lindsay E Fernández-Rhodes**: data interpretation, writing – review & editing

**Olga Garcia-Bedoya**: data interpretation, writing – review & editing

**Qibin Qi**: investigation, data interpretation, writing – review & editing

**Bharat Thyagarajan**: investigation, data acquisition, funding acquisition, data interpretation, writing – review & editing

**Wassim Tarraf**: data interpretation, writing – review & editing

**Tao Wang**: data interpretation, writing – review & editing

**Robert Kaplan**: investigation, data acquisition, funding acquisition, data interpretation, writing – review & editing

**Carmen Isasi**: conceptualization, investigation, data acquisition, funding acquisition, project administration, supervision, data interpretation, writing – original draft, writing – review & editing

**Non-genetic component of height as a surrogate marker for childhood socioeconomic position and its association with cardiovascular and brain health: results from HCHS/SOL**

